# Association of Genetic Variant in MC4R (rs17782313) Gene with Obesity and Other Health Behaviors among Selected Bangladeshi Population: A Population-based Case-control Study

**DOI:** 10.1101/2023.11.14.23298533

**Authors:** Nusrat Jahan Jhily, Lincon Chandra Shill, Dilara Akter Supti, Md Adnan Munim, Rumana Rashid, Roksana Yeasmin, Mohammad Rahanur Alam

## Abstract

**Background and Aims:** This study aim to investigate the influence of a gene polymorphism (rs17782313) on obesity-related traits and biochemical parameters in the Bangladeshi population and the association between this polymorphism and lifestyle-related risk factors.

**Methods:** The study was carried out with 320 samples of which 160 were obese and 160 were healthy people. BMI, MUAC, waist and hip circumference, systolic and diastolic blood pressure, lipid profile, and other socio-demographic and anthropometric profile were accounted for to assess the metabolic properties which are associated with obesity. The tetra-primer Amplified Refractory Mutation System Polymerase chain reaction (ARMS-PCR) was used to genotype rs17782313 in the MC4R gene by using the isolated DNA from collected peripheral blood from the selected sample.

**Result:** The prevalence of hypertension, heart disease, and diabetes were significantly higher among the case group (p<.05) as compared with healthy people. BMI, waist circumference, and hip circumference were significantly higher among people carrying minor allele C (p<.05). We also found a significant difference in dominant (*CC vs. CT+TT*), co-dominant (*CC vs. CT and TT*), and recessive (*TT vs. CC+CT*) model between case and control group (p<.05), which may indicate that rs17782313 in MC4R significantly predict obesity.

**Conclusion:** In conclusion, this study shows the significant association of rs17782313 in MC4R with obesity and obesity-related other health problems. The study has to be conducted further in a broad population to establish a strong association.

## Introduction

Obesity is fetching a crucial public health problem globally which contributes to several preventable metabolic diseases like diabetes, hypertension, cardiovascular disease, and several types of cancers [1]. World Health Organization has defined obesity as a widespread problem because of its histrionic increasing number of overweight and obese people worldwide [2]. According to World Health Organization, the prevalence of overweight and obesity has increased more than doubled between 1980 and 2014, and among them, about 600 million people have obesity [3], and the number increased to >650 million in the year 2016 [4].

This continuous increase in obesity at an alarming rate estimates that the prevalence will rise by around 7.5% by 2025 [5]. A worrying trend has emerged in Bangladesh’s nutritional landscape in recent years: the double burden of malnutrition, comprised of rising obesity and pervasive under-nutrition. At present, about 17% of people are overweight or obese adults in Bangladesh, among which just 4% are obese [6]. The rate of severe chronic health conditions in Bangladesh has risen slowly, and death attributable to chronic conditions increased from 8% in 1986 to 68% in 2006 [7]. Traditionally, infectious diseases and undernutrition were major public concerns in the country, and little attention was given to overweight and obesity by public health officials [8].

Genetic factors are responsible for 40%–70% of the variation in human obesity [9]. An increased risk for obesity has been linked to the MC4R (melanocortin-4 receptor) gene variants, though this association varies by population. [10-12]. The MC4R gene codes for the melanocortin-4 receptor, which is involved in appetite and energy balance regulation [13]. It plays a role in the regulation of appetite and satiety in the brain, influencing feelings of hunger and fullness. The function of the melanocortin-4 receptor may be altered in individuals with specific MC4R polymorphisms, resulting in disruptions in appetite regulation. These disruptions can cause an increase in appetite, a decrease in satiety, and a propensity to consume more food, thereby contributing to weight gain and obesity [14]. Recent studies have linked a common variant (rs17782313) in the MC4R gene’s first intron to an increased risk of obesity and a higher body mass index. Those who inherit the rs17782313 risk allele are more likely to be overweight than those who do not carry the variant [15-18].

Although obesity is a growing public health problem, the underlying genetic factors have received relatively little attention in Bangladesh. The recent study is held with the aim of the identification of dynamic association of genetic variant between manifested MC4R gene SNP rs17782313 and obesity and obesity-related other health behaviors including BMI, biochemical profile, and other lifestyle factors in a selected Bangladeshi population.

## Methods and Materials

This case-control study was conducted in different districts of Bangladesh. But for convenience, we have collected data from the nearest districts like Noakhali, Cumilla, Chandpur, Feni, Dhaka, Lakshmipur, etc. The data were collected from 320 samples among which 160 were cases and 160 were controls. The participants with a BMI of 18.5-24.9 kg/m^2^ were defined as control (healthy) people and the participants with BMI ≥24.9 kg/m^2^ were defined as the case (obese) people [19]. The sample size for this case-control study was calculated using the EPIENFO program (http://www.cdc.gov/epiinfo) [20]. We had considered the ratio between the control and case to be 1:1, Odd Ratio of 1.66 [3] (according to a genetic association study on the Indian population [21]), the percentage of exposure among controls is 60%, among case is 71.3% and 95% confidence interval ratio.

### Inclusion and Exclusion Criteria

Participants aged below 18 years and over 70 years of age were excluded from the study and also the participants with missing values like height, weight, etc. were excluded from the study. During the survey time, the data weren’t collected from pregnant women. The samples were selected with those participants only who matched the inclusion criteria and the data collection process was carried out with the samples and recorded in verbal consent.

### Data collection procedure

We collected both demographic and behavioral information by using a well-structured questionnaire in which the demographic data included anthropometric, socioeconomic, and other personal information and the behavioral information were their dietary habit, physical activities, duration of sleeping, etc.

### Anthropometric analysis

Anthropometric analysis was done by measuring the BMI, hip circumference, and waist circumference when the subject was with light clothing and barefoot. BMI was measured by body weight in kg divided by height in meter square (m^2^) and obesity was determined when individual BMI was ≥24kg/m^2^ and when BMI was 18.5-23.9 kg/m^2^. Hip circumference was measured by wrapping the tape around the individual’s hip at the widest part and the number was then recorded. Waist circumference was measured by wrapping a tape around the waist of individuals and the number was then recorded. The waist-to-hip ratio was then calculated by dividing the waist size by hip size by using a calculator and according to WHO, abdominal obesity was determined by a waist-to-hip ratio of at least 0.90 in men and 0.85 in women.

### Blood sample collection and serum preparation

3ml of venous blood have been collected into a glass container from both the case and control sample. The blood was then kept at room temperature for 1 hour and allowed to clot. After forming a blood clot then the blood sample was centrifuged at 3000 rpm for 20 minutes to separate the serum from the blood cell and stored in a freezer at -20^0^C.

### Biochemical Analysis

The biochemical analysis started with the blood sample collection and serum preparation. Total cholesterol, Triglycerides, HDL-C, and LDL-C were included in the biochemical analysis. Total cholesterol measurement in serum involved the use of three enzymes including cholesterol esterase (CE), cholesterol oxidase (CO), and peroxidase (POD). In the presence of the former, the mixture of ADPS’ and 4-amino antipyrine (4-AA) are condensed by hydrogen peroxide to form a quinone imine dye proportional to the cholesterol concentration in the sample [22]. Triglyceride was measured based on the method of enzymatic hydrolysis of serum or plasma triglyceride to glycerol and free fatty acids (FFA) by lipoprotein lipase (LPL). A red chromogen is produced by the peroxidase (POD) proportional to the concentration of triglyceride in the sample [19]. HDL-Cholesterol was measured by using the method of selective precipitation of apolipoprotein B-containing lipoproteins and residual cholesterol remaining in the supernatant [23]. LDL-Cholesterol was measured by using the Fredwald Equation [23, 24].

### Genotyping

For genomic analysis, DNA extraction kit (Favorgen, Taiwan) was used to isolate DNA. Amplified Refractory Mutation System Polymerase Chain Reaction (ARMS-PCR) was used for genotyping of MC4R single nucleotide polymorphism (SNP) rs17782313 [20]. Tetra-primers were used for the detection of MC4R SNP rs17782313 which were designed by using original Oligo7 software. DNA bands were observed by using UV-transilluminator (UV star, USA) and photographs were taken by using a Gel doc machine (Alpha-imager, USA) attached to a computer, and bands were analyzed. Supplementary Table 1 showed the properties of the primer used in this study.

### Statistical Analysis

The statistical analyses were done using the Statistical Package for Social Science (SPSS). Percentages were used to describe the risk factors for obesity. Means and standard deviations (S.D.) were used for continuous variables and to summarize categorical data both the number and proportion for all the sociodemographic, behavioral, anthropometric, and clinical and biochemical parameters of the study were used. Independent-sample t-tests (for continuous variables), logistic regression, and Pearson’s chi-square test (for categorical variables) were done to perform an intergroup comparison to examine the difference in the covariates between high-risk and not high-risk participants. Analysis of variance (ANOVA) was used to examine the association of MC4R rs17782313 with demographic, biochemical, and BMI.

## Results

### Basic Characteristic features of case and control groups

The demographic, anthropometric, and biochemical profiles of both case and control groups were presented in **Table 1**. Among 320 samples 160 were cases and 160 were in the control group. The anthropometric data showed the mean differences in BMI, MUAC, hip and waist circumference, waist to hip circumference ratio were statistically significant among the case and control groups (p-value<.05). Blood pressure was also higher in obese people than in normal people (p<.05). The biochemical profile showed a significantly higher mean value in the case group than normal included total cholesterol (mg/dl), triglyceride (mg/dl) and LDL-C which were 236.87±37.67, 212.49±34.73, 131.02±41.30 in cases and 206.44±35.27, 198.16±24.22 and 115.18± 15.88 in healthy people respectively (p<.001). The higher biochemical value in the case group indicates a higher risk of having cardiovascular diseases to them. But the mean difference of HDL-C was not statistically significant in both case and control groups (X^2^ = 1.122; p = 0.289).

**Table 1:**
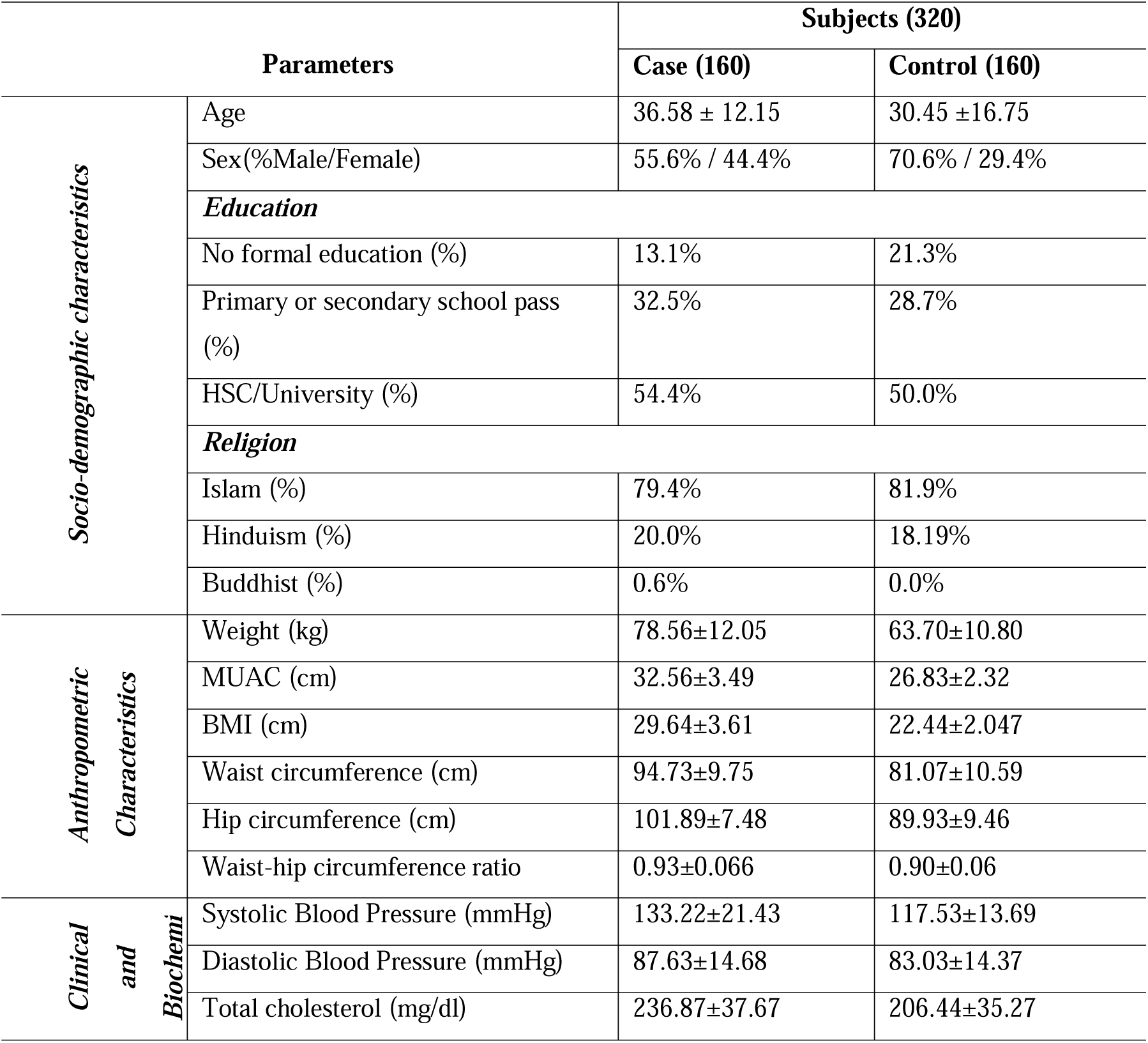
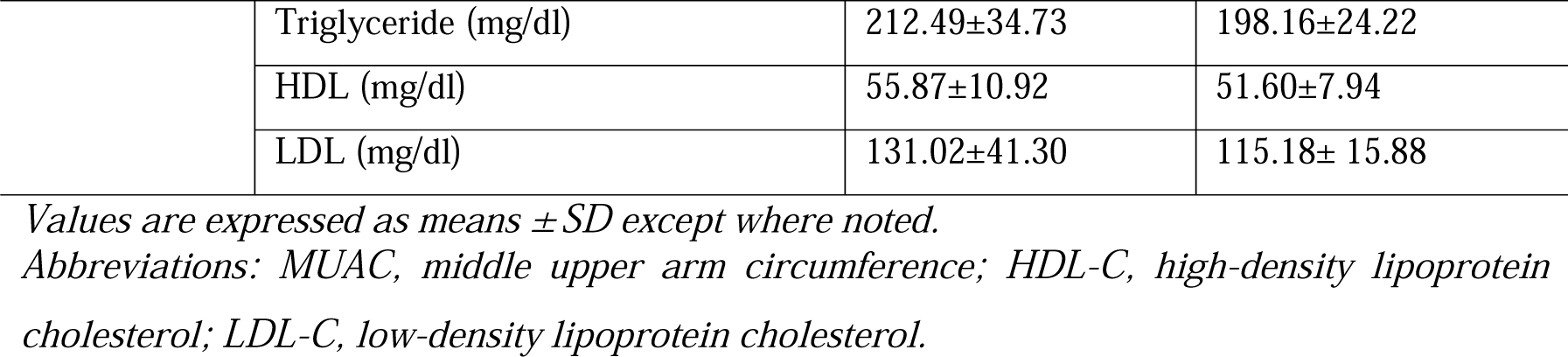
The frequency of sociodemographic, anthropometric, and clinical and biochemical properties compared among obese and healthy people.

### Degenerative disease prevalence

The degenerative disease prevalence among both healthy and obese people was shown in **Table 2**. Among the diseases heart disease, hypertension, diabetes, and kidney diseases were significantly higher in obese people than in normal (p<0.001). The prevalence of arthritis was also higher in the case group but not statistically significant (x^2^ = 2.119; p = 0.224).

**Table 2:** Correlation among various types of degenerative diseases with obesity in both case and control groups.

| Variables |  | Subjects (n=320) |  |  |  |
| --- | --- | --- | --- | --- | --- |
| | | Case (n=160) | Control (n=160) | $\chi^2$ | P-value |
| Heart disease | Yes | 81(84.4%) | 15(15.6%) | 64.821 | 0.000 |
|  | No | 79(35.3%) | 145(64.7%) |  |  |
| Hypertension | Yes | 91(80.5%) | 22(19.5%) | 65.133 | 0.000 |
|  | No | 79(33.3%) | 138(66.7%) |  |  |
| Diabetes | Yes | 96(84.2%) | 18(15.8%) | 82.902 | 0.000 |
|  | No | 64(31.1%) | 142(68.9%) |  |  |
| Arthritis | Yes | 12(66.7%) | 6(33.3%) | 2.119 | 0.224 |
|  | No | 148(49.0%) | 154(51.1%) |  |  |
| Gout | Yes | 29(67.4%) | 14(32.5%) | 6.045 | 0.021 |
|  | No | 131(47.1%) | 146(52.9%) |  |  |
| Kidney disease | Yes | 36(70.0%) | 9(30.0%) | 18.851 | 0.000 |
|  | No | 124(45.1%) | 151(54.9%) |  |  |

### Association of degenerative disease risk factors with obesity among both obese and healthy people

The univariate regression provided the relationship between the obesity and obesity-related degenerative disease risk factors in the case and control group was given in **Table 3**. The analysis showed that systolic and diastolic blood pressure, total cholesterol, triglyceride, and low-density lipoprotein were higher in the case group than in the control group (p<.05), which considered the higher risks of having non-communicable diseases in obese people. The good cholesterol or high-density lipoprotein level was slightly lower in obese people than in healthy, but the data was not statistically significant (p>.05).

**Table 3:** Association of clinical and biochemical profiles of respondents with obesity.

| Variables | Univariate | p-value |
| --- | --- | --- |
|  | OR(95%CI) |  |
| Systolic blood pressure (mmHg) | 0.885 (0.860-0.912) | **<.005** |
| Diastolic blood pressure (mmHg) | 0.893 (0.868-0.919) | **<.001** |
| Total cholesterol (mg/dL) | 0.971 (0.962-0.980) | **.000** |
| Total triglycerides (mg/dL) | 0.980 (0.971-0.989) | **<.005** |
| HDL-C (mg/dL) | 0.988 (0.964-1.013) | .351 |
| LDL-C (mg/dL) | 0.976 (0.968-0.984) | **<.005** |

### Analysis of MC4R variant

Three-point mutations in prominent candidate MC4R gene were genotyped by tetra-primer ARMS PCR-based methodology. PCR fragments were generated as per expectations for all the loci. All the PCR products were well resolved and sized by agarose gel electrophoresis, allowing easy identification of different genotypes. Heterozygotes and homozygotes were unambiguously assigned from the gel profile. The size of DNA fragments amplified with these four primers was between 184 to 351bp and was suitable for separation on 2% agarose gels **(Figure 1)**.

**Figure 1:**
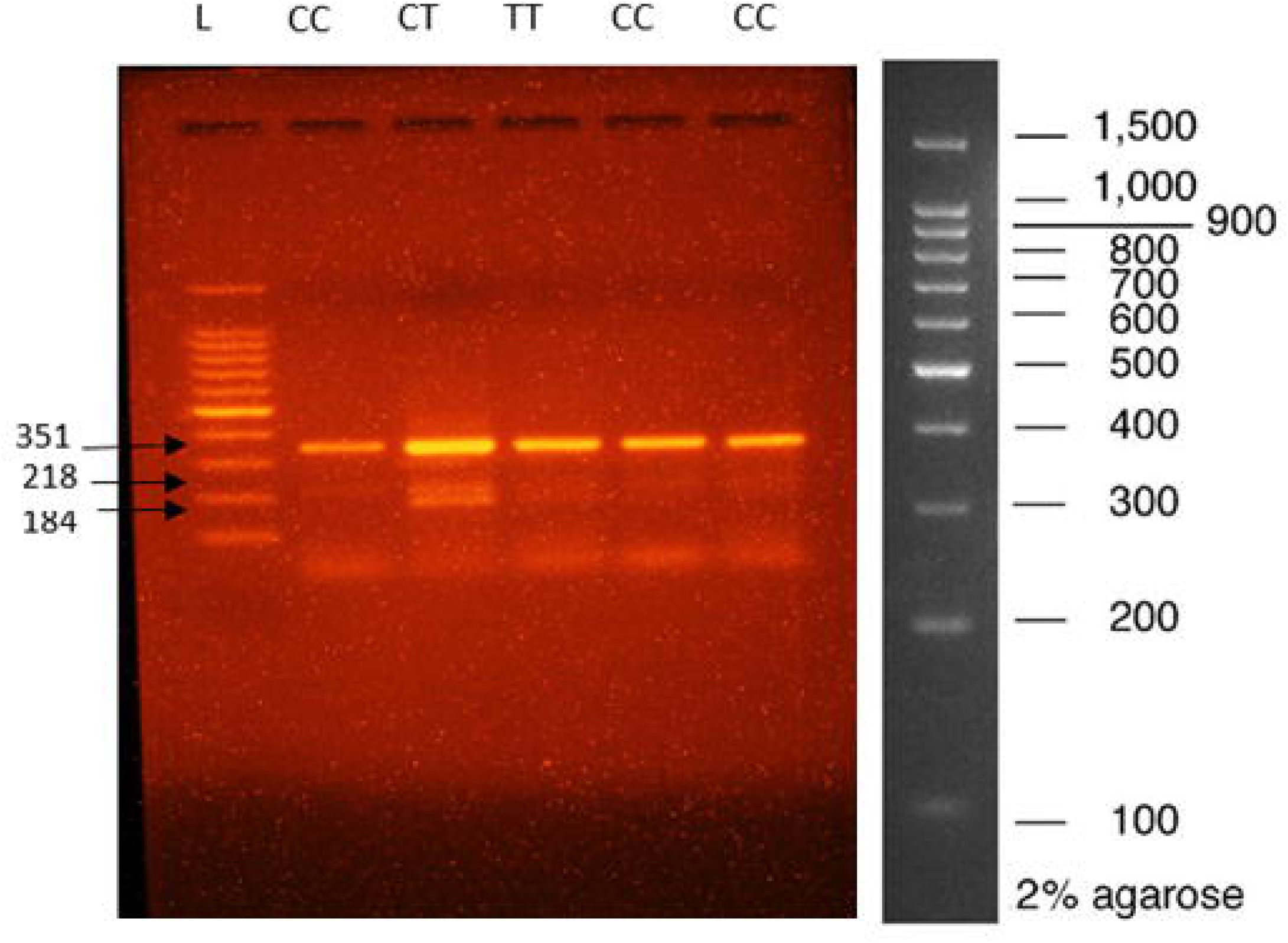
Agarose gel image of ARMS PCR product with 1 kb plus full ranger 100 bp DNA ladder (cat.11800).

In this figure, ARMS PCR analysis of **MC4R (rs17782313)** SNP with Full Ranger 100 bp DNA ladder molecular weight marker (Thermo Scientific), showed that in lanes 2,5,6 CC homozygous mutant and lane 3 CT heterozygous mutant whereas lane 4 TT homozygous wild type. Here lane 1 was used for the ladder to detect the genotypes.

In **Table 4** the association of MC4R gene polymorphism with case and control is presented. In the Co-dominant model, we found that people with homozygous wild-type genotype TT are 0.177 times more likely to have obesity (p<0.005), whereas people with homozygous mutant genotype CC are statistically significantly 5.643 times more likely to have obesity (p<0.005) than people with TT genotype. People with heterozygous mutant genotype CT are shown 3.415 times more likely to have obesity compared with people with TT genotype, which is also statistically significant (p<.005). By looking at the Dominant model, we found people with the CT+TT genotype are 0.306 times more likely to have obesity compared with people with the CC genotype (p<.005). On the other hand, in the recessive model, people with the CC+CT genotype have been found as 4.598 times more risks of becoming obese compared with people with TT genotype, which is statistically significant (p<.005).

**Table 4:**
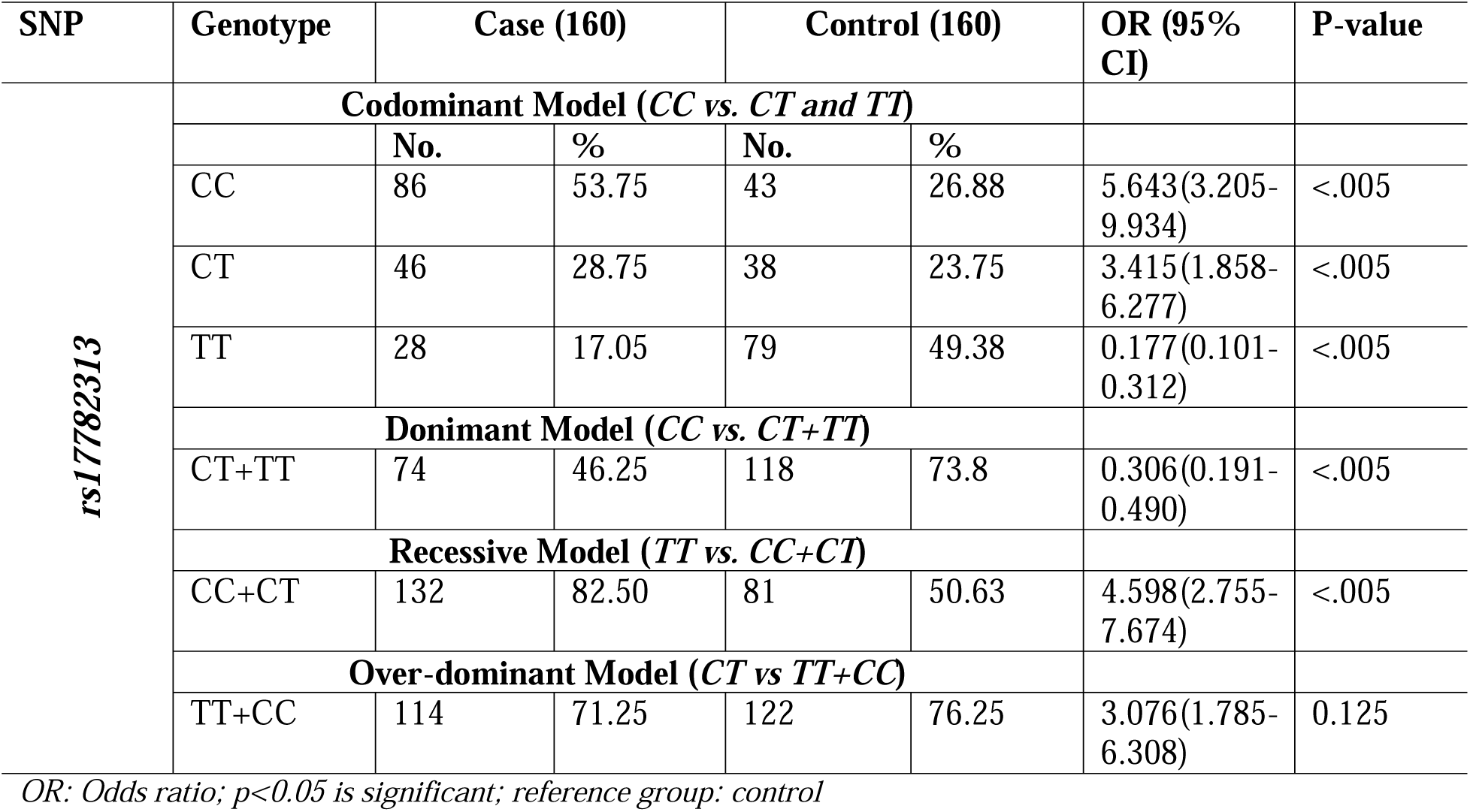
Genotypes of MC4R gene polymorphism model analysis.

Turning to the genotype model analysis in more detail, we found there people with the TT+CC genotype have 3.076 times more risks of having obesity compared with people with the CT genotype in the Over-dominant model. But this result was not statistically significant (p>.05).

### Comparison of MC4R gene variant with demographic, anthropometric, and biochemical characteristics in both case and control groups

**Table 5** provided the association of genetic variants in MC4R (rs17782313) with the socio-demographic, anthropometric, and clinical properties of obese and healthy people. The presence of homozygous allele CC and heterozygous allele CT were associated with higher BMI, waist circumference, and hip circumference (p<.05) which are closely related to obesity. The data also showed that the presence of at least one C allele in obese people was associated with increased blood pressure, total cholesterol, triglycerides, LDL-C, etc., but there was no significant association (p>.05). Looking at the dataset of the control group we can see that people with homozygous allele CC and heterozygous allele CT have slightly higher values of systolic and diastolic blood pressure, total cholesterol, triglyceride, LDL-C, and lower value of HDL-C than people with homozygous allele TT. This is like the presence of at least one C allele is responsible having obesity and obesity-like degenerative diseases, although the data did not show a statistically significant association (p>.05). Further we can see that the CC allele is associated with higher FCS scores in the control group which is statistically significant (p<.05).

**Table 5:**
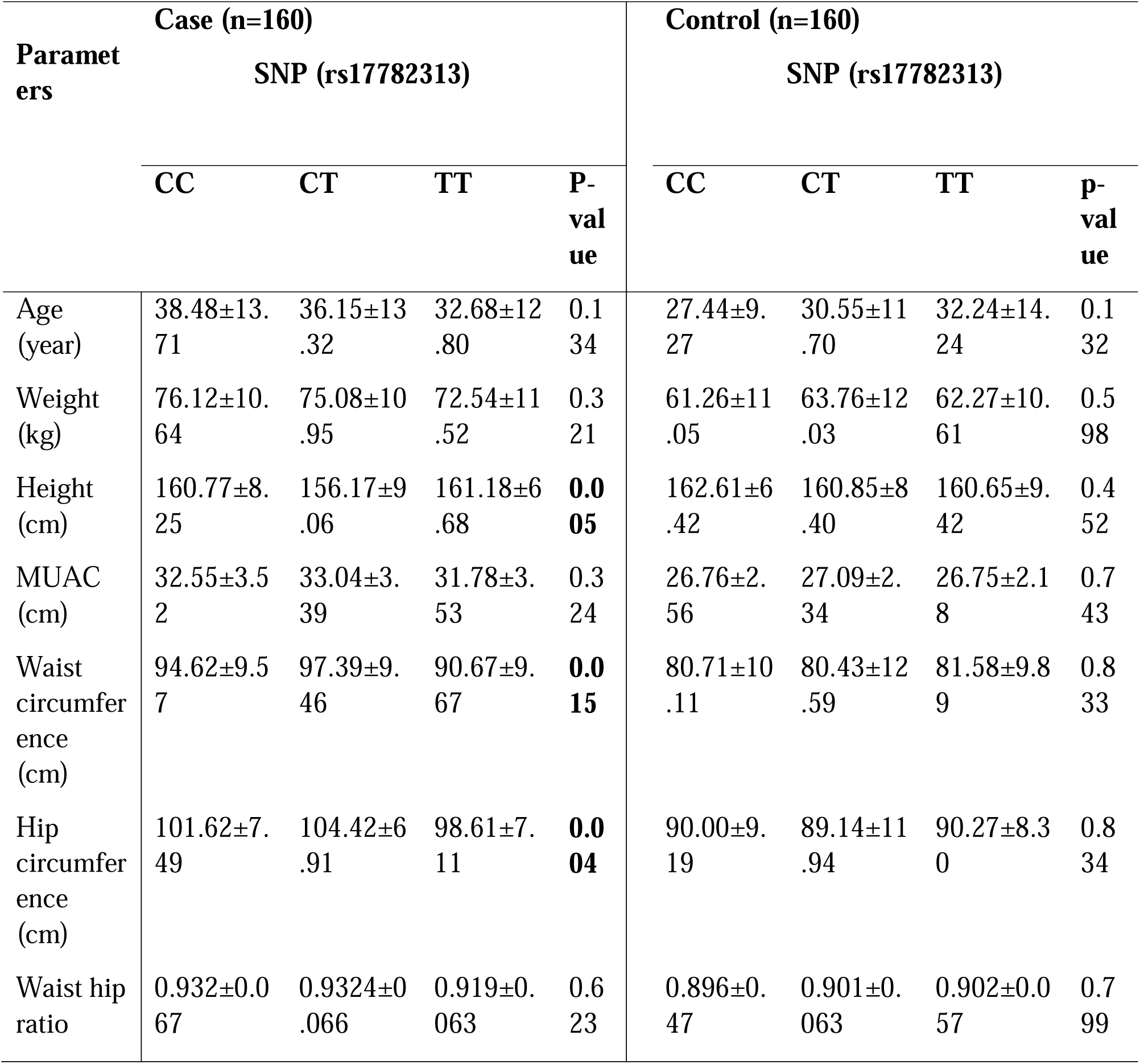
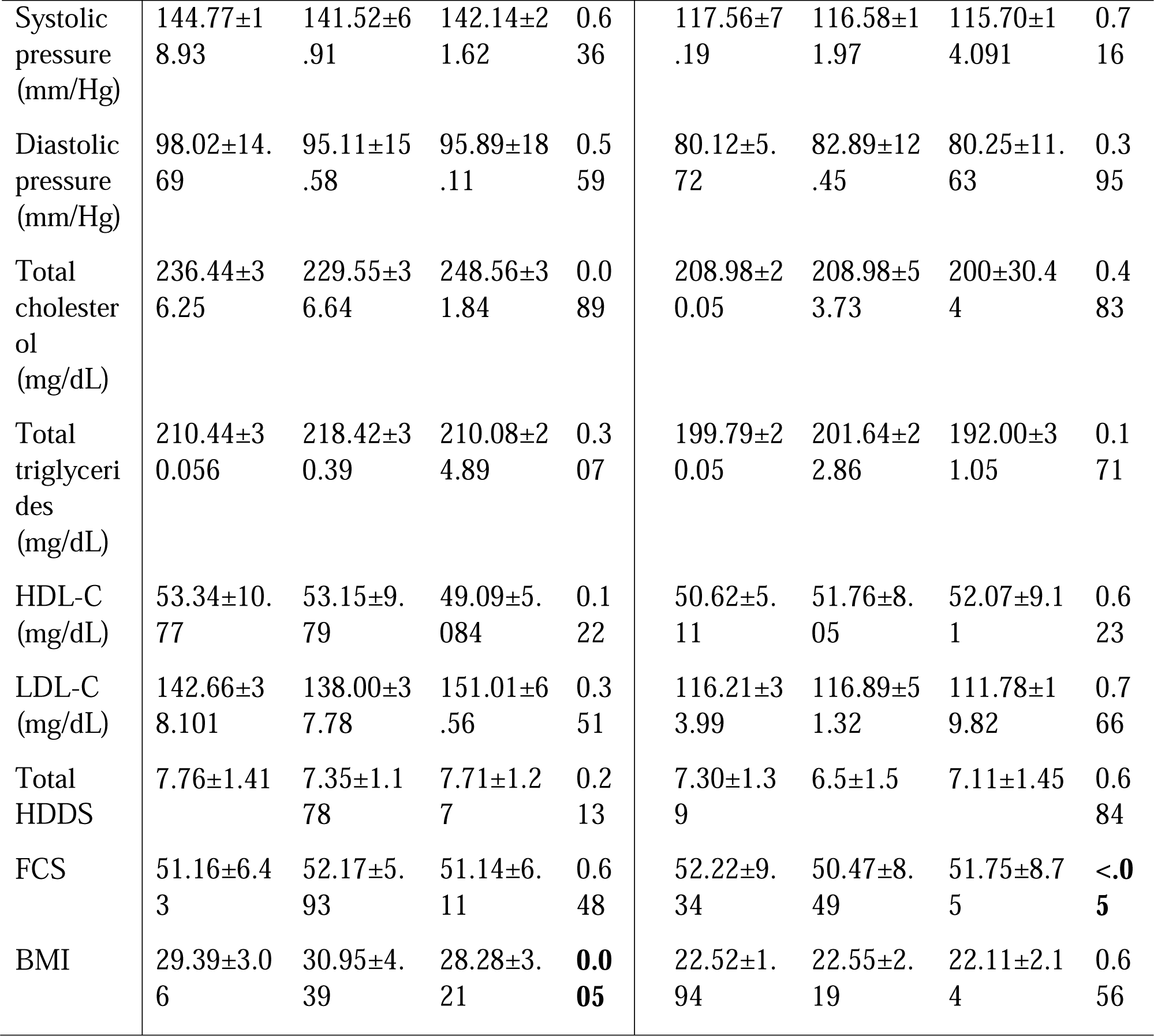
Comparison of demographic, anthropometric, and biochemical parameters among different genotypes of MC4R SNP rs17782313 in healthy and obese populations.

Further, **Table 6** showed the comparison among different genotypes according to the dominant model in both healthy and obese groups. Height, hip-circumference, and BMI were found significantly higher in the case group carrying the CC+CT allele (p<.05). There is no significant association found between genotype and obesity-related other health parameters, biochemical and dietary parameters. However, the highest mean of MUAC, waist-circumference, triglyceride, and LDL-C are found in obese people carrying the CC+CT allele.

**Table 6:** Comparison of demographic, anthropometric, and biochemical parameters among different genotypes of MC4R rs17782313 in the healthy and obese populations (dominant model)

| Parameters | Case (n=160)<br>SNP (rs17782313) |  |  | Control (n=160)<br>SNP (rs17782313) |  |  |
| --- | --- | --- | --- | --- | --- | --- |
|  | TT | CC+CT | p-value | TT | CC+CT | p-value |
| Age (year) | 32.68±12.80 | 34.82±13.65 | 0.085 | 32.24±14.24 | 31.79±13.07 | 0.325 |
| Weight (kg) | 72.54±11.52 | 73.95±11.44 | 0.225 | 62.27±10.61 | 63.11±11.37 | 0.635 |
| Height (cm) | 161.18±6.68 | 158.88±7.97 | **0.035** | 160.65±9.42 | 160.92±8.97 | 0.438 |
| MUAC (cm) | 31.78±3.53 | 32.4±3.65 | 0.276 | 26.75±2.18 | 26.98±2.35 | 0.175 |
| Waist circumference (cm) | 90.67±9.67 | 94.03±9.76 | 0.125 | 81.58±9.89 | 81.05±11.34 | 0.850 |
| Hip circumference (cm) | 98.61±7.11 | 101.72±7.21 | **0.01** | 90.27±8.30 | 89.71±10.23 | 0.775 |
| Waist hip ratio | 0.919±0.063 | 0.926±0.0647 | 0.742 | 0.902±0.057 | 0.9017±0.12 | 0.825 |
| Systolic pressure (mm/Hg) | 142.14±21.62 | 141.93±14.37 | 0.823 | 115.70±14.091 | 116.25±13.05 | 0.628 |
| Diastolic pressure (mm/Hg) | 95.89±18.11 | 95.75±16.94 | 0.635 | 80.25±11.63 | 81.64±12.15 | 0.678 |
| Total cholesterol (mg/dL) | 248.56±31.84 | 239.25±34.44 | 0.147 | 200.00±30.44 | 204.54±42.17 | 0.680 |
| Total triglycerides (mg/dL) | 210.08±24.89 | 214.35±27.74 | 0.427 | 192.00±31.05 | 196.94±27.02 | 0.750 |
| HDL-C (mg/dL) | 49.09±5.084 | 51.32±7.54 | 0.236 | 52.07±9.11 | 51.98±8.64 | 0.225 |
| LDL-C (mg/dL) | 151.01±6.56 | 144.61±22.27 | 0.462 | 111.78±19.82 | 114.43±35.62 | 0.498 |
| Total HDDS | 7.71±1.27 | 7.73±1.32 | 0.343 | 7.11±1.45 | 6.7±1.53 | 0.105 |
| FCS | 51.14±6.11 | 51.84±6.32 | 0.678 | 51.75±8.75 | 51.21±8.73 | 0.213 |
| BMI | 28.28±3.21 | 29.72±3.65 | **0.02** | 22.11±2.14 | 22.42±2.13 | 0.876 |

*MUAC (middle-upper arm circumference); HDL-C (high-density lipoprotein cholesterol); LDL-C (low-density lipoprotein cholesterol); HDDS (household dietary diversity score); FCS (food consumption score); BMI (body mass index); p<0.05 is significant*

## Discussion

Obesity is becoming a worldwide epidemic in both developed and developing countries. In Bangladesh there is not enough study held between MC4R with obesity and obesity-related parameters and also data are very limited. Therefore, it is required to organize further assessment between obesity and other biochemical and genetic features to find a clear association between these. Because obesity is comprised of several multifactorial pathogeneses, it is essential to use an approach to analyze the association of the MC4R gene with obesity, including considerations of socioeconomic status, lifestyle factors, and obesity-related measurements. We conducted this study with two groups’ cases (obese people) and control (healthy people) which were selected based on their BMI. We found from our study, BMI, MUAC, waist and hip circumference, and waist-to-hip circumference ratio was statistically significantly higher among the case group compared with the healthy group. Body mass index (BMI) and Waist-circumference were good indicators for assessing overweight and obesity and also in some cases another indicator known as Waist-to-hip (WHR) ratio worked as an alternative indicator to measure obesity [25, 26].

In this study, we found that significantly higher prevalence of obesity-related multi-morbidity among obese people than among non-obese. A higher prevalence of diabetes, hypertension, and heart disease is found in our recent study in obese people than in healthy. Studies also found that higher prevalence of hypertension and type 2 diabetes are closely related to obesity [27, 28]. The higher mean of blood pressure, total cholesterol, triglyceride, and LDL-C are found in the case group as compared with the control group in this study, which was significantly associated with a higher prevalence of non-communicable diseases and it was in line with a Brazilian study where it was indicated that higher level of total cholesterol, triglycerides, and LDL-C were the major risk factors of having heart disease in overweight and obese people [29]. A study organized by da Fonseca et. al. also found a higher value of anthropometric and biochemical parameters in obese people than in normal [30]. Another study conducted by Luying Gao et. al. found that higher mean of systolic and diastolic blood pressure, total cholesterol, and LDL-C which was closely related to obesity and obesity-related degenerative diseases [31].

Our present study found the association between MC4R gene SNP rs17782313 is related to an increased level of blood pressure, total cholesterol, triglycerides, and LDL-C which is mostly similar to other studies [32, 33]. This study also found that people carrying at least one C allele were at the condition of higher BMI and at increased risk of obesity and obesity-related metabolic disorders, which is similar to many other studies [34]. A genome-wide association study which was conducted in Caucasian people found some genetic variants associated with obesity and among them, rs17782313 of MC4R was the second strongest SNP associated with higher BMI and increased risk of having obesity [35]. The SNP rs17782313 was significantly associated with an increased risk of obesity and obesity-connected anthropometric parameters mostly in females found in one gender-specific study held in Pakistan [33].

In this study, the presence of homozygous allele CC and heterozygous allele CT was significantly associated with increased BMI, waist circumference, and hip circumference, which is similar to a Korean study that identified the variant rs17782313 was associated with higher BMI in people with minor allele C [36]. According to a case-control study by Le Thi Tuyet et al., in Vietnam, the anthropometric indices of the case group carrying the CC allele were higher than the TT allele and also the anthropometric indices were higher as compared with the control group [34]. Another study conducted on Chinese people and Tatar women also found a strong association between MC4R rs17782313 with increased BMI in subjects carrying CC and CT alleles [37, 38]. The higher mean of total cholesterol, triglycerides, and LDL-C were identified in obese people carrying minor allele C, although the association was not significant in our recent study. A Multi-Institutional Collaborative Cohort (J-MICC) study conducted on Japanese people showed a significant association of high serum triglyceride levels in people carrying minor allele C with rs17782313. The study also acclaimed that rs17782313 was highly associated with a higher level of serum triglyceride in Asian people carrying at least one C allele in their genotype [39]. We examined the association of MC4R rs17782313 and obesity with four different models, among them we found significant associations in the co-dominant, dominant, and recessive models. Another case-control study also significantly identified the correlation between rs17782313 and obesity in dominant and co-dominant models [40].

Our study is the first genetic study in Bangladesh showing the association of genetic variant in Melanocortin-4 Receptor (MC4R) with obesity and obesity-related anthropometric, biochemical, dietary, and metabolic traits and this is the strongest point of the study. However, there were a lot of limitations in our study. We didn’t establish any significant association with dietary patterns. Further metabolic status with obesity and rs17782313 was not also strongly examined. The risk factors were not also included. There may be some misconception happened if the participants give any biased information about their disease status. It should be needed to examine in the laboratory. Ultimately, the study was conducted based on laboratory measures of a blood sample which may have over-accounted the biochemical profile.

## Conclusion

In conclusion, our study confidently established that being overweight and obese is significantly associated with different anthropometric, biochemical, and degenerative diseases. It is also evident that the MC4R is closely associated with obesity and an increased risk of heart disease and hypertension in Bangladeshi people. Obesity prevalence is increasing rapidly which is also associated with many other non-communicable diseases. Therefore, it is of great importance that we need to examine the risk factors of these diseases and the biochemical profile of obese people. Further, it should be needed to raise awareness to prevent obesity and its complications. To prevent and treat obesity, obese adults should be recommended for early detection and examining health problems. A repeated study also needs to be conducted to inquire into the exact mechanism of MC4R and obesity and obesity-related other parameters.

## Supporting information

Supplementary Table 1 showed the properties of the primer used in this study.

## Data Availability

Data will be made available upon reasonable request.

## Notes

**Funding Statement:** This work was supported by the NSTU research grant 2020-2021 by the Research cell of Noakhali Science and Technology University (NSTU/RC-FTNS/T-21/71).

### Competing Interest Statement

The authors have declared no competing interest.

### Funding Statement

This work was supported by the NSTU research grant 2020-2021 by the Research cell of Noakhali Science and Technology University (NSTU/RC-FTNS/T-21/71).

### Author Declarations

The ethical approval was obtained from the institutional ethics committee of the Noakhali Science and Technology University.

## References

1. Norman, J.E., The adverse effects of obesity on reproduction. Reproduction, 2010. 140(3): p. 343.

2. Organization, W.H., Obesity: preventing and managing the global epidemic. 2000: World Health Organization.

3. Dankyau, M., et al., Prevalence and correlates of obesity and overweight in healthcare workers at a tertiary hospital. Journal of Medicine in the Tropics, 2016. 18(2): p. 55.

4. Organization, W.H., World Health Organization Obesity, and Overweight.

5. Collaboration, N. and N.R.F. Collaboration, Trends in the adult body-mass index in 200 countries from 1975 to 2014: a pooled analysis of 1698 population-based measurement studies with 19· 2 million participants. Lancet, 2016. 387(10026): p. 1377–96.

6. Arthur, M., Institute for health metrics and Evaluation. Nursing Standard (2014+), 2014. 28(42): p. 32.

7. Swinburn, B.A., et al., The global obesity pandemic: shaped by global drivers and local environments. The Lancet, 2011. 378(9793): p. 804-814.

8. Chowdhury, M.A.B., M.M. Adnan, and M.Z. Hassan, Trends, prevalence and risk factors of overweight and obesity among women of reproductive age in Bangladesh: a pooled analysis of five national cross-sectional surveys. BMJ Open, 2018. 8(7): p. e018468.

9. Barsh, G.S., I.S. Farooqi, and S. O’rahilly, Genetics of body-weight regulation. Nature, 2000. 404(6778): p. 644-651.

10. Frayling, T.M., et al., A common variant in the FTO gene is associated with body mass index and predisposes to childhood and adult obesity. Science, 2007. 316(5826): p. 889-894.

11. Dina, C., et al., Variation in FTO contributes to childhood obesity and severe adult obesity. Nature Genetics, 2007. 39(6): p. 724–726.

12. Loos, R. and C. Bouchard, FTO: the first gene contributing to common forms of human obesity. Obesity Reviews, 2008. 9(3): p. 246–250.

13. Krashes, M.J., B.B. Lowell, and A.S. Garfield, Melanocortin-4 receptor–regulated energy homeostasis. Nature neuroscience, 2016. 19(2): p. 206–219.

14. Garfield, A.S., et al., A neural basis for melanocortin-4 receptor–regulated appetite. Nature neuroscience, 2015. 18(6): p. 863–871.

15. Huang, W., Y. Sun, and J. Sun, Combined effects of FTO rs9939609 and MC4R rs17782313 on obesity and BMI in Chinese Han populations. Endocrine, 2011. 39: p. 69–74.

16. Lurie, G., et al., The obesity-associated polymorphisms FTO rs9939609 and MC4R rs17782313 and endometrial cancer risk in non-Hispanic white women. PLoS One, 2011. 6(2): p. e16756.

17. Marcadenti, A., et al., Effects of FTO RS9939906 and MC4R RS17782313 on obesity, type 2 diabetes mellitus and blood pressure in patients with hypertension. Cardiovascular diabetology, 2013. 12: p. 1–8.

18. Yilmaz, Z., et al., Association between MC4R rs17782313 polymorphism and overeating behaviors. International journal of obesity, 2015. 39(1): p. 114–120.

19. Dean, A., OpenEpi: open source epidemiologic statistics for public health, version 2.3. 1. http://www.openepi.com, 2010.

20. Dean, A., K. Sullivan, and M. Soe, OpenEpi: open source epidemiologic statistics for public health, version. 2014.

21. Prakash, J., et al., Association of FTO rs9939609 SNP with obesity and obesity-associated phenotypes in a north Indian population. Oman medical journal, 2016. 31(2): p. 99.

22. Meiattini, F., et al., The 4-hydroxybenzoate/4-aminophenazone chromogenic system used in the enzymic determination of serum cholesterol. Clinical chemistry, 1978. 24(12): p. 2161–2165.

23. Grove, T.H., Effect of reagent pH on determination of high-density lipoprotein cholesterol by precipitation with sodium phosphotungstate-magnesium. Clinical chemistry, 1979. 25(4): p. 560–564.

24. Warnick, G.R., et al., Estimating low-density lipoprotein cholesterol by the Friedewald equation is adequate for classifying patients on the basis of nationally recommended cutpoints. Clinical chemistry, 1990. 36(1): p. 15–19.

25. Yang, F., et al., Receiver-operating characteristic analyses of body mass index, waist circumference and waist-to-hip ratio for obesity: Screening in young adults in central south of China. Clinical Nutrition, 2006. 25(6): p. 1030–1039.

26. Ahmad, N., et al., Abdominal obesity indicators: Waist circumference or waist-to-hip ratio in Malaysian adults population. International journal of preventive medicine, 2016. 7.

27. Xi, B., et al., Common polymorphism near the MC4R gene is associated with type 2 diabetes: data from a meta-analysis of 123,373 individuals. Diabetologia, 2012. 55: p. 2660–2666.

28. Da Fonseca, A.C.P., et al., Identification of the MC4R start lost mutation in a morbidly obese Brazilian patient. Diabetes, Metabolic Syndrome and Obesity: Targets and Therapy, 2019: p. 257–266.

29. de Almeida, C.A., et al., Abdominal circumference as an indicator of clinical and laboratory parameters associated with obesity in children and adolescents: comparison between two reference tables. Jornal de Pediatria, 2007. 83: p. 181–185.

30. Da Fonseca, A.C.P., et al., Genetic variants in the activation of the brown-like adipocyte pathway and the risk for severe obesity. Obesity Facts, 2020. 2(2): p. 130–143.

31. Gao, L., et al., MC4R single nucleotide polymorphisms were associated with metabolically healthy and unhealthy obesity in Chinese northern Han populations. International journal of endocrinology, 2019. 2019.

32. Xi, B., et al., Association between common polymorphism near the MC4R gene and obesity risk: a systematic review and meta-analysis. 2012.

33. Rana, S., S. Rahmani, and S. Mirza, MC4R variant rs17782313 and manifestation of obese phenotype in Pakistani females. RSC advances, 2018. 8(30): p. 16957–16972.

34. Le Thi, T. and Q.B. Tran, Associations of Single Nucleotide Polymorphism rs17782313 in Melanocortin 4 Receptor Gene with Anthropometric Indices in Normal and Obesity Primary School Children in Hanoi. VNU Journal of Science: Medical and Pharmaceutical Sciences, 2018. 34(2).

35. den Hoed, M., et al., Genetic susceptibility to obesity and related traits in childhood and adolescence: influence of loci identified by genome-wide association studies. Diabetes, 2010. 59(11): p. 2980–2988.

36. Scherag, A., et al., Two new Loci for body-weight regulation identified in a joint analysis of genome-wide association studies for early-onset extreme obesity in French and german study groups. PLoS genetics, 2010. 6(4): p. e1000916.

37. Kochetova, O., et al., Association of polymorphic variants of FTO and MC4R genes with obesity in a Tatar population. Genetika, 2015. 51(2): p. 248–255.

38. Hong, J., et al., Genetic susceptibility, birth weight and obesity risk in young Chinese. International journal of obesity, 2013. 37(5): p. 673–677.

39. Katsuura-Kamano, S., et al., A polymorphism near MC4R gene (rs17782313) is associated with serum triglyceride levels in the general Japanese population: the J-MICC Study. Endocrine, 2014. 47: p. 81–89.

40. Abd Ali, A.H., The common gene MC4R rs17782313 polymorphism associated with obesity: A meta-analysis. Human Gene, 2022: p. 201035.

