## Supplementary Table 1 showed the properties of the primer used in this study. for "Association of Genetic Variant in MC4R (rs17782313) Gene with Obesity and Other Health Behaviors among Selected Bangladeshi Population: A Population-based Case-control Study"

**Consent:** Do you want to participate in this study after knowing the pros and cons of the study?

Yes	□ /	No	□

A. **Personal information**

| 1. Name of the participant: |
| --- |
| 2. Father/Husband/Legal guardian’s name: |
| 3. Date of birth: |
| 4. Address and contact information: |
| 5. Religion:   Answer key: 1. Islam; 2. Hinduism; 3. Buddhism; 4. Christianism; 5. Atheism; 6. Other (----------------) |
| 6. Sex:   Answer key: 1. Male; 2. Female; 3. Third sex |
| 7. Education level:   Answer key: 1. Illiterate; 2. Literate; 3. Passed Primary school; 4. Passed Junior school; 9. Passed SSC; 10. Passed HSC; 11. University/College graduate |

B. **Anthropometric information**

| 8. Weight (kg): |
| --- |
| 9. Height (cm): |
| 10. BMI: |
| 11. MUAC (cm): |
| 12. Waist circumferencea (cm): |
| 13. Hip circumferencea (cm): |
| 14. Head circumferenceb (cm): |
| 15. Systolic blood pressure (mmHg) |
| 16. Diastolic blood pressure (mmHg) |

C. **Biochemical information:**

| 17. Total cholesterol (mg/dL) |
| --- |
| 18. Total triglycerides (mg/dL) |
| 19. HDL-C (mg/dL) |
| 20. Total cholesterol (mg/dL) |

D. **Dietary intake-related information**

**Food Consumption Score (FCS) and Household Dietary Diversity Score (HDDS):** Only for adult participants. *(Show Flash)*

|  | **Food** | **FCS**  **Eaten in past 7 days**  ***If 0 days, do not specify the main source.*** | **HDDS**  **Consumed in the past 24hrs?** |
| --- | --- | --- | --- |
|  | **Note: We have to determine whether fish and milk consumption was only in small quantities.** | **How many days over the last 7 days, did most members of your household (50% +) eat the following food items?** | **Did most of your household eat/consume the following foods yesterday? (yes=1, No=0)** |
| 1. | **Cereals, grains, roots and tubers (FCSstap):** Rice, pasta, bread, sorghum, millet, maize, potato, yam, cassava, white sweet potato |  |  |
| *If 0 skip to question 2* | | |  |
| 1.1. | **Cereals, grains (HDDSStapCer):** rice, pasta, bread, sorghum, millet, maize | \| |  |
| 1.2. | **Roots and tubers (HDDSStapRoot):** potato, yam, cassava, white sweet potato |  |  |
| 2. | **Pulses/legumes/nuts (FCSPulse/HDDSPulse):** beans, cowpeas, peanuts, lentils, nut, soy, pigeon pea and / or other nuts |  |  |
| 3. | **Milk and other dairy products (FCSDairy/HDDSDairy)**: fresh milk / sour, yogurt, cheese, other dairy products  (Exclude margarine / butter or small amounts of milk for tea / coffee) |  |  |
| 4. | **Meat, fish and eggs (FCSpr):** goat, beef, chicken, pork, blood, fish, including canned tuna, escargot, and / or other seafood, eggs (meat and fish consumed in large quantities and not as a condiment) |  |  |
| *If 0 skip to question 5* | | |  |
| 4.1 | **Flesh meat (FCSPrMeatF/HDDSPrMeatF):** beef, pork, lamb, goat, rabbit, chicken, duck, other birds, insects |  |  |
| 4.2 | **Organ meat (FCSPrMeatO/HDDSPrMeatO):** liver, kidney, heart and / or other organ meats |  |  |
| 4.3 | **Fish/shellfish (FCSPrFish/HDDSPrFish):** fish, including canned tuna, escargot, and / or other seafood (fish in large quantities and not as a condiment) |  |  |
| 4.4 | **Eggs (FCSPrEggs/HDDSPrEgg)** |  |  |
| **5.** | **Vegetables and leaves (FCSVeg/HDDSVeg):** spinach, onion, tomatoes, carrots, peppers, green beans, lettuce, etc |  |  |
| If 0 skip to question 6 | | |  |
| 5.1 | **Orange vegetables (vegetables rich in Vitamin A) (FCSVegOrg):** carrot, red pepper, pumpkin, orange sweet potatoes, |  |  |
| 5.2 | **Green leafy vegetables (FCSVegGre):** spinach, broccoli, amaranth and / or other dark green leaves, cassava leaves |  |  |
| **6.** | **Fruits (FCSFruit/HDDSFruit):** banana, apple, lemon, mango, papaya, apricot, peach, etc |  |  |
| *If 0* *skip to question 7* | | |  |
| 6.1 | **Orange fruits (Fruits rich in Vitamin A) (FCSFruitOrg):** mango, papaya, apricot, peach |  |  |
| **7.** | **Oil / fat / butter (FCSFat/HDDSFat):** vegetable oil, palm oil, shea butter, margarine, other fats / oil |  |  |
| **8.** | **Sugar, or sweet (FCSSugar/HDDSSugar):** sugar, honey, jam, cakes, candy, cookies, pastries, cakes and other sweet (sugary drinks) |  |  |
| **9.** | **Condiments / Spices (FCSCond/HDDSCond):** tea, coffee / cocoa, salt, garlic, spices, yeast / baking powder, lanwin, tomato / sauce, meat or fish as a condiment, condiments including small amount of milk / tea coffee. |  |  |
